# Genomic surveillance of human metapneumovirus in the United States, 2010 – 2025

**DOI:** 10.64898/2026.01.18.26344359

**Authors:** Emily E. Bendall, William J Fitzsimmons, Weronika Damek Valvano, Rachel Truscon, Wesley H. Self, Natasha Halasa, James D. Chappell, Yuwei Zhu, Basmah Safdar, Adit Ginde, Ithan D. Peltan, Manjusha Gaglani, Cristie Columbus, Nathan I. Shapiro, Kevin W. Gibbs, David N. Hager, Matthew Prekker, Amira Mohamed, Nicholas J. Johnson, Jay S. Steingrub, Akram Khan, Abhijit Duggal, Jennifer G. Wilson, Nida Qadir, Laurence W. Busse, Jennie H. Kwon, Matthew C. Exline, Ivana A. Vaughn, Jarrod M. Mosier, Estelle S. Harris, Fatimah S. Dawood, Kevin C. Ma, Diya Surie, Arnold S. Monto, Emily T. Martin, Adam S. Lauring

## Abstract

Human metapneumovirus (HMPV) is a significant cause of acute respiratory illness in both children and adults, yet its genomic epidemiology remains understudied compared to other respiratory viruses. Here, we report an expanded genomic surveillance of HMPV in the United States, utilizing 325 newly sequenced samples from two cohorts: the household-based HIVE study (2010–2022) in Michigan and the multicenter IVY network (2022–2025) of hospitalized adults. Our analyses revealed the continued predominance of the A2.2.2 clade and little geographic structure in the US. Genomic diversity is highest in the glycoprotein (G); we identified a shift from variants bearing the 180nt duplication to ones with a 111nt duplication. Phylogenetic analyses supported the duplication-deletion model for the origin of the duplications. The conserved fusion (F) protein shows limited antigenic variation and low rates of nonsynonymous substitutions, suggesting stability in epitopes targeted by vaccine candidates. These findings underscore the utility of enhanced genomic surveillance for understanding HMPV evolution and informing vaccine development.

**Summary:** Genomic surveillance of HMPV in the United States over a 15 year period reveals extensive genetic diversity and more rapid evolution in the G protein compared to the F protein, which is targeted by vaccines under development.

## Introduction

Initially identified in 2001, human metapneumovirus (HMPV) is recognized as a leading cause of mild and severe respiratory infections in young children^1–4^. HMPV is comparable to respiratory syncytial virus (RSV) in its overall incidence, symptom frequency, and acute respiratory illness (ARI) burden among younger and older adults^5,6^. However, there is increasing recognition that the elderly and immunocompromised are at higher risk for severe infections requiring hospitalization^7^. This increased recognition of severe HMPV-associated illness has coincided with increased, but still limited, genomic surveillance of HMPV subtype, clade, and strain diversity^8,9^.

HMPV is a negative-sense, single stranded RNA virus that shares many characteristics with RSV. Of the 9 proteins encoded by the HMPV genome, the three surface proteins – fusion (F), glycoprotein (G), and small hydrophobic (SH) proteins – can be targeted by host antibodies. The G protein is an important virulence factor that affects aspects of both innate and adaptive immunity^10,11^ but is weakly immunogenic^12^. Like the RSV F protein, HMPV F is more conserved and is targeted by vaccines currently in development^13–15^. Indeed, the F proteins are sufficiently similar between the two viruses that some antibodies are specific for both^16,17^.

HMPV is genetically diverse with two antigenically distinct lineages (A and B) that cocirculate globally ^18,19^. Both HMPV-A and HMPV-B are further divided into two sublineages: A1, A2 and B1, B2, respectively. The A2 sublineage has given rise into multiple clades: A2.1, A2.2, A2.2.1, and A2.2.2 ^20^. Most of the genetic diversity across subtypes lies within G ^21,22^, which is also highly variable within subtypes. Within A2.2.2, two duplications in G have occurred; a 111nt duplication first detected in 2017 and 180nt duplication first detected in 2015^23,24^. Originally, the duplications were thought to be independent events^24^. More recently a new hypothesis, the duplication-deletion model, has been proposed, whereby the 180nt duplication occurred, followed by a partial deletion leading to the 111nt lineage. The data thus far are inconclusive.

The SARS-CoV-2 pandemic advanced recognition of the public health benefits of genomic surveillance further. Genomic surveillance of SARS-CoV-2, influenza viruses, RSV, mpox, and other agents has been critical for identifying emerging lineages, preventing their spread, and updating vaccines to ensure high effectiveness^25^. Until recently, however, there has been relatively limited whole genome sequencing of HMPV. Prior to January 2025 there were 513 hMPV-A and 456 hMPV-B genomes available in NCBI Genbank. Furthermore, most of the recent (post-2020) hMPV genomes within the United States come from Washington state^9^. Public health would greatly benefit from broader genomic characterization and surveillance of HMPV, especially with vaccine candidates in clinical trials.

Here we expand upon HMPV genomic surveillance in two important ways. First, we substantially expand the geographic range of recent HMPV genomes in the United States. We sequenced contemporary samples (2022-2025) from the Investigating Respiratory Viruses in the Acutely Ill (IVY) network, which consists of 26 medical centers in 20 U.S states. Second, we sequenced historical samples (2010-2022) from Michigan and the Household Influenza Vaccine Effectiveness (HIVE) study. This expanded sampling across the US and over a longer period allowed us to elucidate important evolutionary dynamics of HMPV.

## Methods

Samples were obtained from two cohorts: The Household Influenza Vaccine Effectiveness (HIVE) study and Investigating Respiratory Viruses in the Acutely Ill (IVY) network.

HIVE is a prospective, longitudinal cohort of households with children maintained for surveillance of symptomatic acute respiratory illnesses and based in Ann Arbor, Michigan. Participants were instructed to report illnesses meeting a standard case definition: the onset of 2 or more symptoms of ARI from any member in the household. Nasal and/or throat samples were collected at the time of symptom onset. Study design, laboratory methods, and consent procedures have been described previously^26,27^. Written informed consent was obtained from adults. Parents or legal guardians of minor children provided written informed consent on behalf of their children. The study protocols were reviewed and approved by the University of Michigan Institutional Review Board (HUM118900 & HUM198212). We identified all HIVE samples positive for HMPV between 2010 and 2022.

IVY is an inpatient network of adults hospitalized with ARI at 26 medical centers across the US. ARI was defined as presence of at least 1 of the following: fever, cough, dyspnea, chest imaging findings consistent with pneumonia, or hypoxemia. Nasal samples were collected from individuals who had a positive nucleic acid test result for HMPV within 10 days of symptom onset and 3 days of admission. Enrollment occurred between July 1, 2022 and April 30, 2025.

This activity was reviewed by CDC and by the institutional review boards at each participating institution, deemed public health surveillance and not research with a waiver of participant informed consent, and was conducted consistent with applicable federal law and CDC policy (45 CFR part 46.102(l)(2), 21CFR part 56; 42 USC §241(d); 5 USC §552a; 44 USC §3501, et seq).

### Sequencing

All samples with a cycle threshold < 32 were processed for whole genome sequencing. RNA was extracted using the MagMAX viral/pathogen nucleic acid purification kit (ThermoFisher) and a KingFisher Flex instrument. Libraries were prepared using the Illumina Respiratory Virus Enrichment Kit, and sequenced on an Illumina NextSeq 2000 (2x300, P1 chemistry). Fastq files were downsampled to 500,000 reads. Consensus sequences were generated using IRMA^28,29^. Clades were determined by Nextclade^30^. Consensus sequences with > 80% coverage and a “good” Nextclade quality score were used for further analyses.

### Phylogenetic Analysis

Phylogenetic analyses were performed separately for HMPV-A and HMPV-B. Sequences were aligned using MAFFT^31^. IQ-TREE^32^ was used for model selection and inferring the phylogeny. 1000 bootstrap replicates were performed. TreeTime^33^ was used to calibrate a time-based phylogeny and for estimating evolutionary rates. Additionally, separate F and G gene phylogenies were inferred. All phylogenies were visualized using ggtree^34^.

For the F ectodomain, we examined all single nucleotide polymorphisms (SNPs) found in 2 or more individuals. We excluded SNPs in which the mutation was clade defining (i.e., at an internal node giving rise to the clade). We calculated dN/dS using SLAC^35^ for each gene and subtype. We also examined the duplication in G within A2.2.2. A2.2.2 samples were classified as having the 111nt duplication, 180nt duplication, or no duplication. The duplication status was “undetermined” for samples with <85% coverage of G.

## Results

We identified 247 HMPV positive samples from the HIVE community cohort (2010-2022) and 369 HMPV positive samples from the IVY network of hospitalized adults (2022-2025) with quantities sufficient for sequencing. From these 616 samples, we successfully sequenced 325 samples (> 80% genomic coverage) from IVY (n=233) and HIVE (n=92). HIVE samples were mostly from children and IVY was exclusively samples from adults (Table 1). We successfully sequenced samples for every season between 2010/11 and 2024/25, except for 2020/21, when there was little identified circulation of HMPV (Figure 1A). Ninety-six were HMPV-B, 229 were HMPV-A, and 18 states were represented (Figure 1B). A2.2.2 was the dominant clade for HMPV-A across seasons (Figure 1C). For HMPV-B, B1 and B2 circulated in varying levels across seasons. Starting in the 2022-23 season, B2 has continually been dominant (Figure 1C). There was no geographical structuring of clades evident for either HMPV-A or HMPV-B in the nationally representative IVY network (Figure 2).

**Figure 1.**
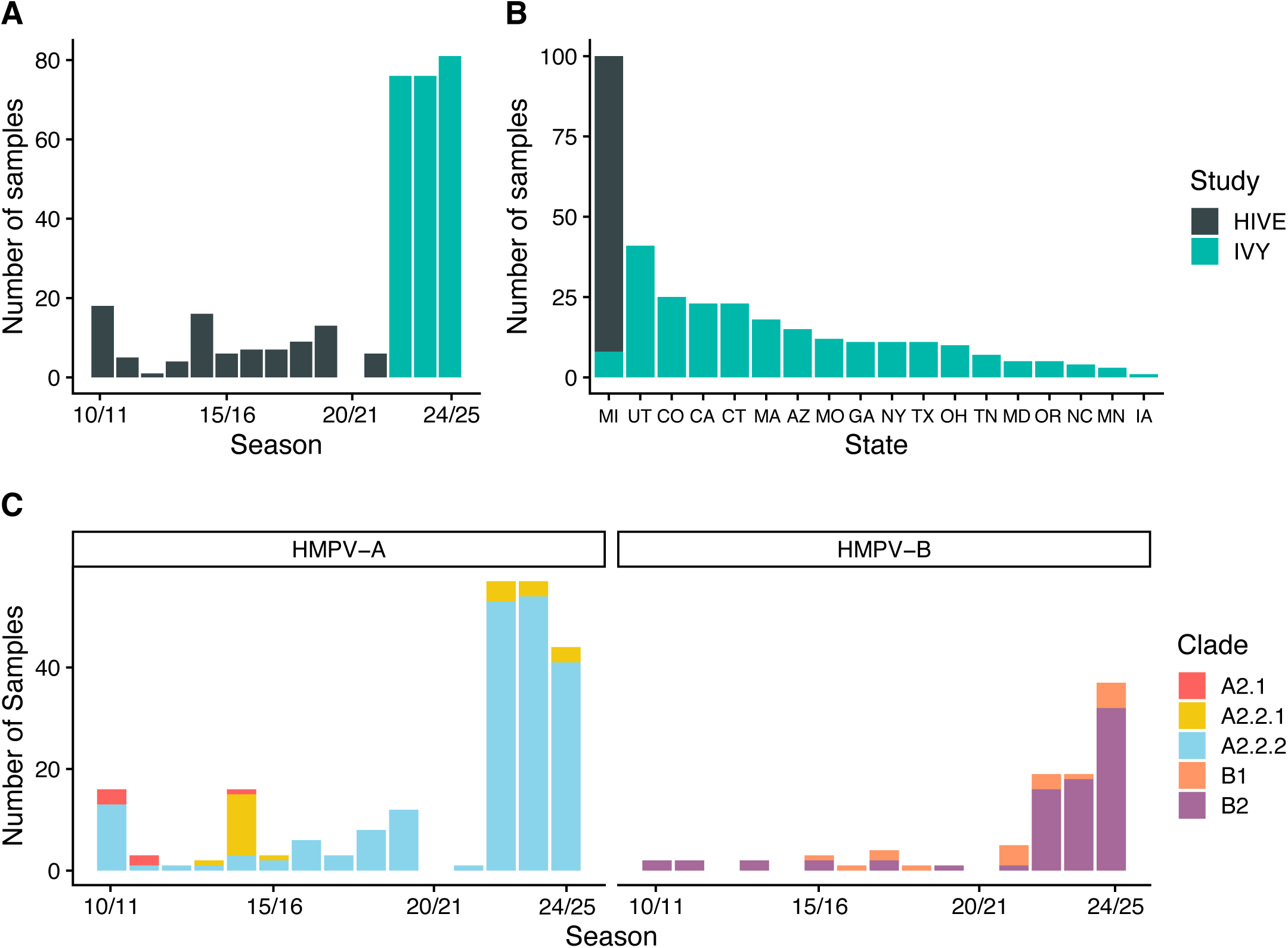
Overview of successfully sequenced samples. A. The number of samples sequenced per season colored by study. Grey is HIVE and teal is IVY. B. The number of samples from each state. C. The Number of HMPV-A and HMPV-B samples per season colored by clade.

**Figure 2.**
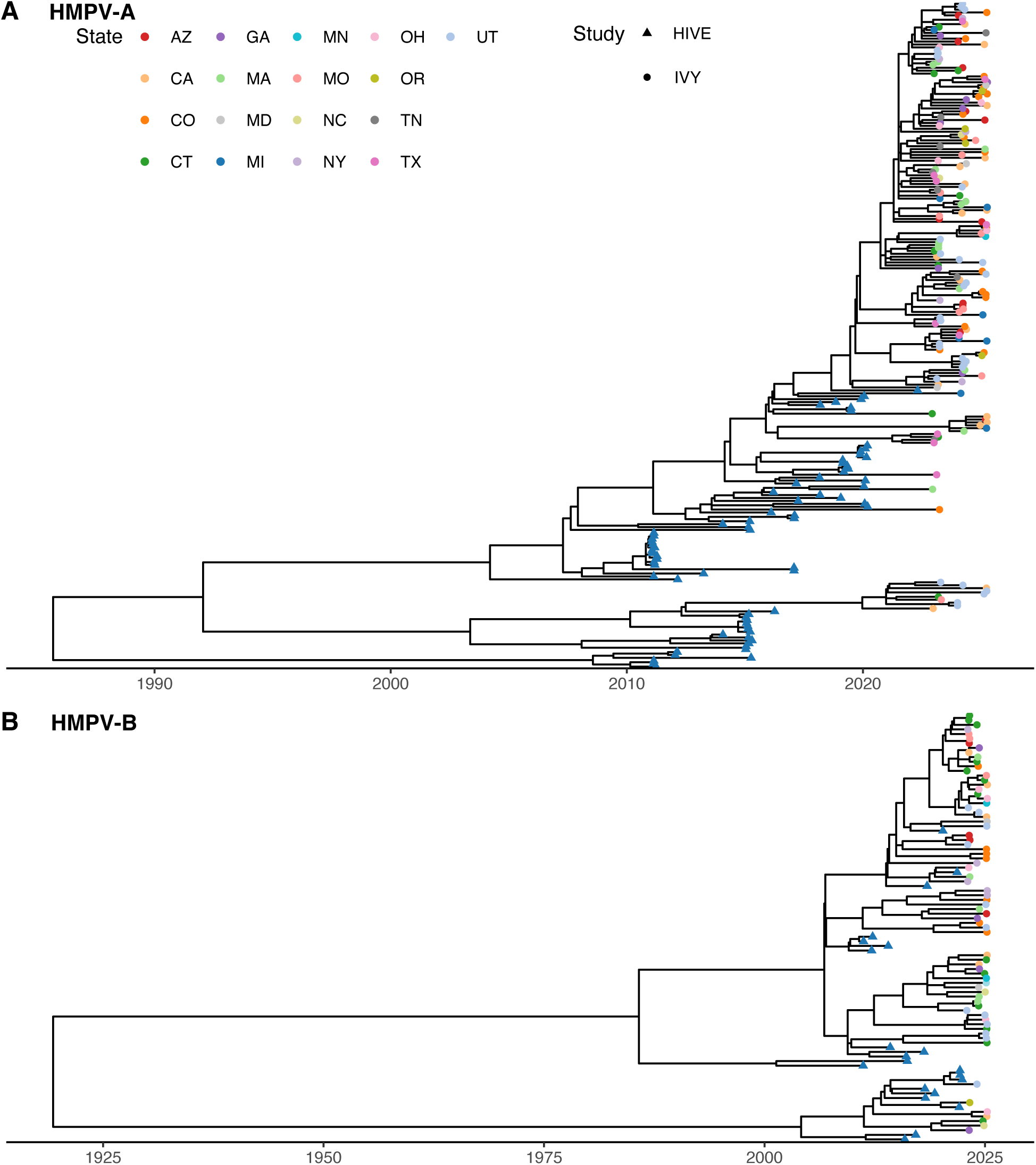
Whole genome phylogeny for (A) HMPV-A and (B) HMPV- B. HMPV-A and HMPV-B lack geographic structure in our sample. Tips are colored by the state, and the tip shape is the study.

**Table 1.** Age of participants with successfully sequenced HMPV by study, United States, 2010–2025, N=325.

|  | HIVE | IVY |
| --- | --- | --- |
|  | n = 92 | n = 233 |
| Age, years | n (%) | n (%) |
| 0-5 | 54 (58.7) | 0 (0) |
| 6-17 | 23 (25.0) | 0 (0) |
| 18-50 | 14 (15.2) | 35 (15.0) |
| 51-65 | 1 (1.1) | 55 (23.6) |
| >65 | 0 (0) | 143 (61.4) |

For HMPV-A, 3 SNPs were identified in the ectodomain of F (Figure 3A). A K171R substitution was found in 3 individuals in epitope ∅, which is immunodominant in RSV but a less prominent epitope in HMPV^36^. The K296R substitution, found in two individuals, is the main determinant of strain related fusion phenotypes and the binding site of a monoclonal antibody (4F11)^37,38^. For HMPV-B, 10 SNPs were identified in the ectodomain of F (Figure 3B). Five of these were in known epitope sites: L36H/Q (site III), K142R (site V), R175I (site ∅), and R229K (site II). Epitopes II, III, and V are immunodominant in HMPV^36^. There was also a SNP at 179, which is a known monoclonal antibody binding site (SAN32-2 Fab)^39^. Like HMPV-A, there was a SNP at position 296, although the amino acid change was different (K296R vs D296N).

**Figure 3.**
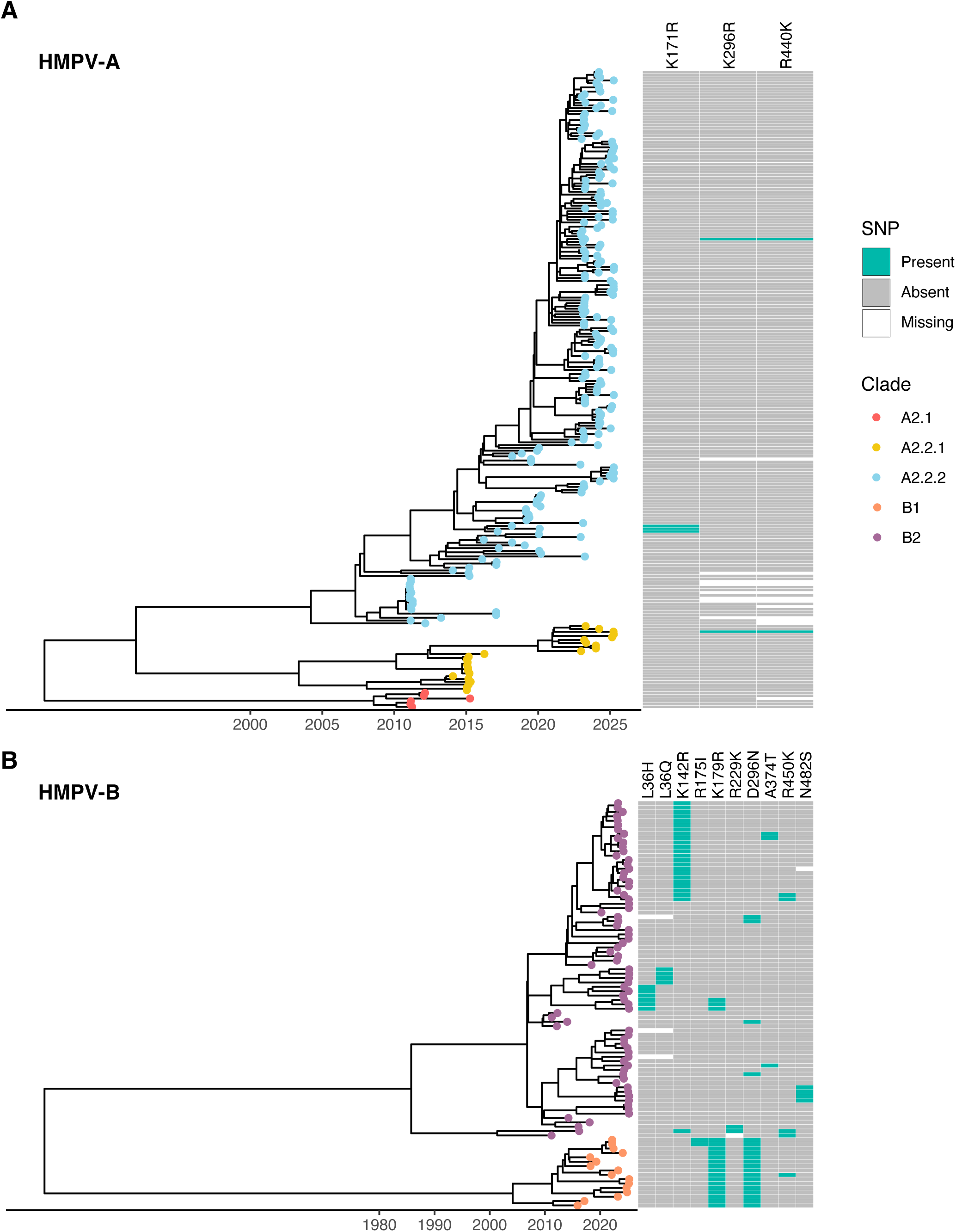
Whole genome phylogeny for (A) HMPV-A and (B) HMPV- B colored by clade with a heatmap of amino acid substitutions in the F ectodomain.

The divergence rate for HMPV-A based on the whole genome was 9.13x10^-4^ (std = 3x10^-5^ ) substitutions/site/year. F had a similar divergence rate of 7.16x10^-4^ substitutions/site/year (std=1x10^-4^). G had a faster divergence rate of 2.85x10^-3^ substitutions/site/year (std= 3x10^-4^). HMPV-B followed the same pattern. The whole genome divergence rate was 7.32x10^-4^ substitutions/site/year (std=3x10^-5^). It was 6.53x10^-4^ substitutions/site/year (std= 1x10^-4^) for F, and 2.12x10^-3^ substitutions/site/year (std= 3x10^-4^) for G. Consistent with divergence rates, G had higher amino acid diversity across the gene than F (Figure S1).

Despite the higher divergence rate and amino acid diversity, dN/dS < 1 for G (Table 2), which is indicative of purifying selection. All genes had a dN/dS < 1, but the value for G was much closer to 1 than the other genes in both HMPV-A and HMPV-B. The higher dN/dS may be due to relaxed purifying selection across the gene or alternatively indicate specific sites under positive selection with the rest of the gene under purifying selection.

**Table 2.** dN/dS for HMPV-A and HMPV-B by gene.

| Gene | HMPV-A | HMPV_B |
| --- | --- | --- |
| N | 0.047 | 0.036 |
| P | 0.115 | 0.106 |
| M | 0.028 | 0.025 |
| F | 0.038 | 0.057 |
| M2-1 | 0.092 | 0.071 |
| M2-2 | 0.268 | 0.074 |
| SH | 0.366 | 0.375 |
| G | 0.652 | 0.757 |
| L | 0.060 | 0.0481 |

Within G, all A2.2.2 samples had a duplication (Figure 4). The 180nt duplication was present from 2010 and onwards (Figure 4A). The 111nt duplication was identified beginning in 2016. The 111nt duplication slowly became dominant within A2.2.2, and for the last two seasons, only the 111nt duplication has circulated. These data support a duplication-deletion model (Figure 4B), as our data resolve that the 111nt clade is a subclade of the 180nt clade, and not a separate monophyletic clade. The duplication-deletion model is also supported by the F and G phylogenies (Figure S2).

**Figure 4.**
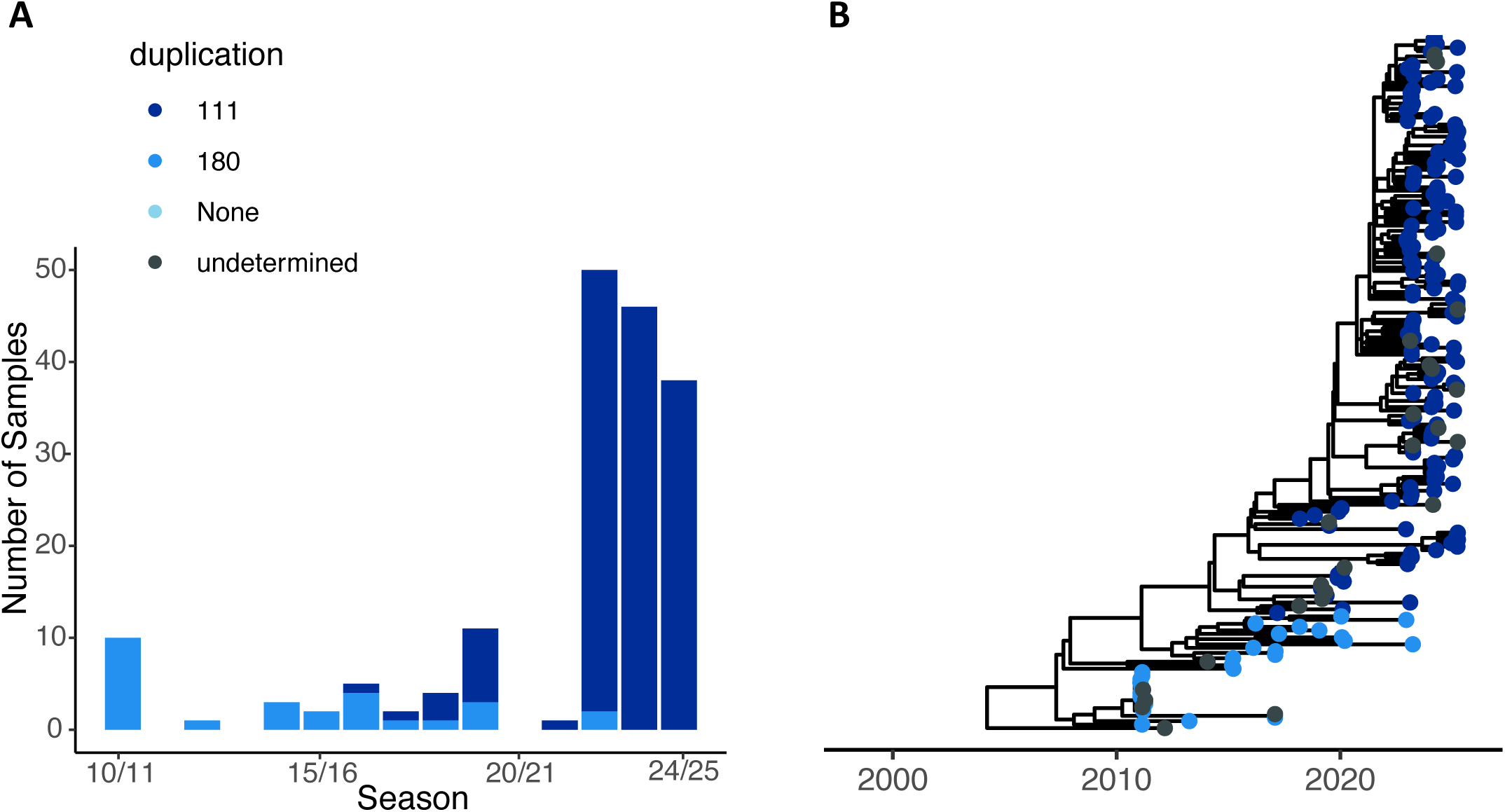
HMPV-A A2.2.2 samples. (A) The number of samples per season with each duplication type. (B) Whole genome phylogeny for (A) HMPV-A A2.2.2 samples. The colors represent the duplication type within G.

## Discussion

The genomic evolution and epidemiology of HMPV have been understudied relative to some other respiratory viruses. Here we addressed this deficiency, increasing the available data in public data repositories by 25% with a single study. Our sample includes individuals across the age and illness severity spectrum, and in recent years, across the United States. The scope of this study provides a clear picture of the clade dynamics of HMPV over the last 15 years and insights into its longer-term evolutionary dynamics.

The patterns we observe in the U.S. are consistent with those observed globally^9,40^. From 2010 to 2015, A2.1, A2.2.1, and A2.2.2 cocirculated. After 2015, A.2.2.2 became predominant, especially the lineage with the 111nt duplication. After 2020, A2.2.2 viruses containing the 111nt duplication has predominated with some co-circulation of A2.2.1. For HMPV-B, B2 has been the predominant strain over the last three years. Additionally, our whole genome divergence rates are similar to previous rates 1.11ll×ll10^−3^ substitutions/site/year vs 9.13x10^-4^ for HMPV-A and 8.93ll×ll10^−4^ substitutions/site/year vs 7.16x10^-4^ for HMPV-B^9^.

Our study resolves the relationship between the 180nt and 111nt duplication. Previously, there were mixed data on whether these duplications represented two separate duplication events, or a duplication followed by a partial deletion. Goya et al. inferred a phylogeny from all whole genome sequences available globally^9^. The resulting phylogeny supported either scenario equally. However, their G-based phylogeny supported the duplication-deletion model. We sequenced an additional 50 A2.2.2 samples collected between 2010 and 2020, which further resolved the phylogeny over the period spanning the duplication events. Our whole genome, F, and G-based phylogenies all supported the duplication deletion model. Given the rapid spread of the 180nt A2.2.2 lineage followed by the 111nt A2.2.2 lineage, the duplication is likely to be advantageous. As proposed in RSV, the duplication in G may cause G to be an immune decoy. With the duplication, G protrudes further from the membrane than F, which potentially prevents antibodies from accessing the F epitopes^41^. The partial deletion in G may represent a selective tradeoff between immune evasion and structural stability.

The evolutionary dynamics of F are very different than those of G. F has a lower divergence rate compared to the genome-wide rate. There are very few SNPs in the ectodomain within clades, and even fewer SNPs in antigenic sites. There are three SNPs in immunodominant epitopes for HMPV-B and none in HMPV-A. Additionally, four of the HMPV-B and all HMPV-A amino acid changes are Arginine to Lysine, or vice versa. Lysine and Arginine are both positively charged basic amino acids, and substitutions are less likely to have significant effects on protein structure. The low number of substitutions in F epitopes is promising for F-based vaccines as it suggests an absence of significant antigenic drift, which would require regular updates to the vaccine. Continued genomic surveillance will be important to ensure this remains true over time.

This study has several strengths. First, our addition of a substantial number of HMPV genomes with linked time and location data allows for better genomic surveillance and greater understanding of HMPV dynamics. Second, retrospectively sequencing earlier samples allowed us to determine the origins of the duplication within G. One of the benefits of sequencing samples from a range of timepoints in a single study is that all the samples were sequenced using the same method and processed with the same bioinformatic pipelines. Duplications in the glycoprotein can easily be missed if not explicitly factored into the analysis. For example, RSV has similar duplications in the RSV-B and RSV-A glycoprotein and aligning samples to references that predate the duplications has resulted in misassembly and failed detection of the duplications^42^. This probably occurs in HMPV as well and can mask the phylogenetic signal of the origin of the duplication when using genomes from a variety of sources. Misassembly of whole genome next generation sequencing may be responsible for previous inconsistencies in the duplication origin. In contrast to the whole genome sequencing, many of the earlier G samples were Sanger sequenced and correctly capture the presence of the duplications.

There are also important limitations to this study. Although we took a multi-site approach, our data is U.S. centric, and there may be regional patterns on a global scale that are not captured in our data. Geographical bias in sampling is a problem for HMPV genomics generally. The majority of HMPV genomes come from the United States, China, and Australia. More geographically diverse sampling is important for prompt identification of emerging lineages and mutations that may impact the effectiveness of future vaccines or antivirals. Next, the use of hospitalized patients in recent years may bias our results from the contemporary samples (IVY) if lineages differ in severity, or if they differ in age groups affected. Importantly, however, our results are consistent with previous sampling, and our data are still likely to identify major patterns.

Our expanded genomic surveillance of HMPV provides important insights into the evolutionary dynamics of this important respiratory pathogen. These data substantially increase available HMPV genomes and enhance resolution of phylogenetic relationships, offering a foundation for ongoing surveillance and vaccine evaluation. There is a continued need for broader global genomic surveillance to capture regional diversity and rapidly identify emerging mutations with potential clinical implications. As HMPV vaccine development progresses, integrating routine whole-genome surveillance will be essential for monitoring viral evolution and informing public health interventions.

## Data Availability

All genomes are available at NCBI GenBank (PRJNA1181811), and raw reads are available at NCBI SRA (PRJNA1304962). All analysis code is available at https://github.com/lauringlab/HMPV_Genomic_Epidemiology.

## Author Contributions

Conceiving and Designing the Study: ASL, ETM, WHS, DS

Acquiring the Data: All authors

Analysis and Interpretation of the Data: EEB, WDV, ASL

Drafting the Manuscript: EEB, ASL

Editing the Manuscript: All authors

Supervising the Research: ASL, WHS

Obtained Funding: ASL, ETM, WHS

The findings and conclusions in this report are those of the authors and do not necessarily represent the official position of the Centers for Disease Control and Prevention.

## Acknowledgements

We thank the participants in the IVY Network and HIVE study. IVY was supported by the CDC (contract 75D30122C14944 to Vanderbilt University Medical Center). HIVE was supported in part with federal funds from the National Institute of Allergy and Infectious Diseases, National Institutes of Health, Department of Health and Human Services (contract 75N93021C00015); and the Centers for Disease Control and Prevention (grants U01IP001034, R01AI097150, R56AI097150, U18IP000170, U01IP000474). Sequencing and analysis were supported in part by the National Institute of Allergy and Infectious Diseases (U19AI181767).

---

**Figure S1.**
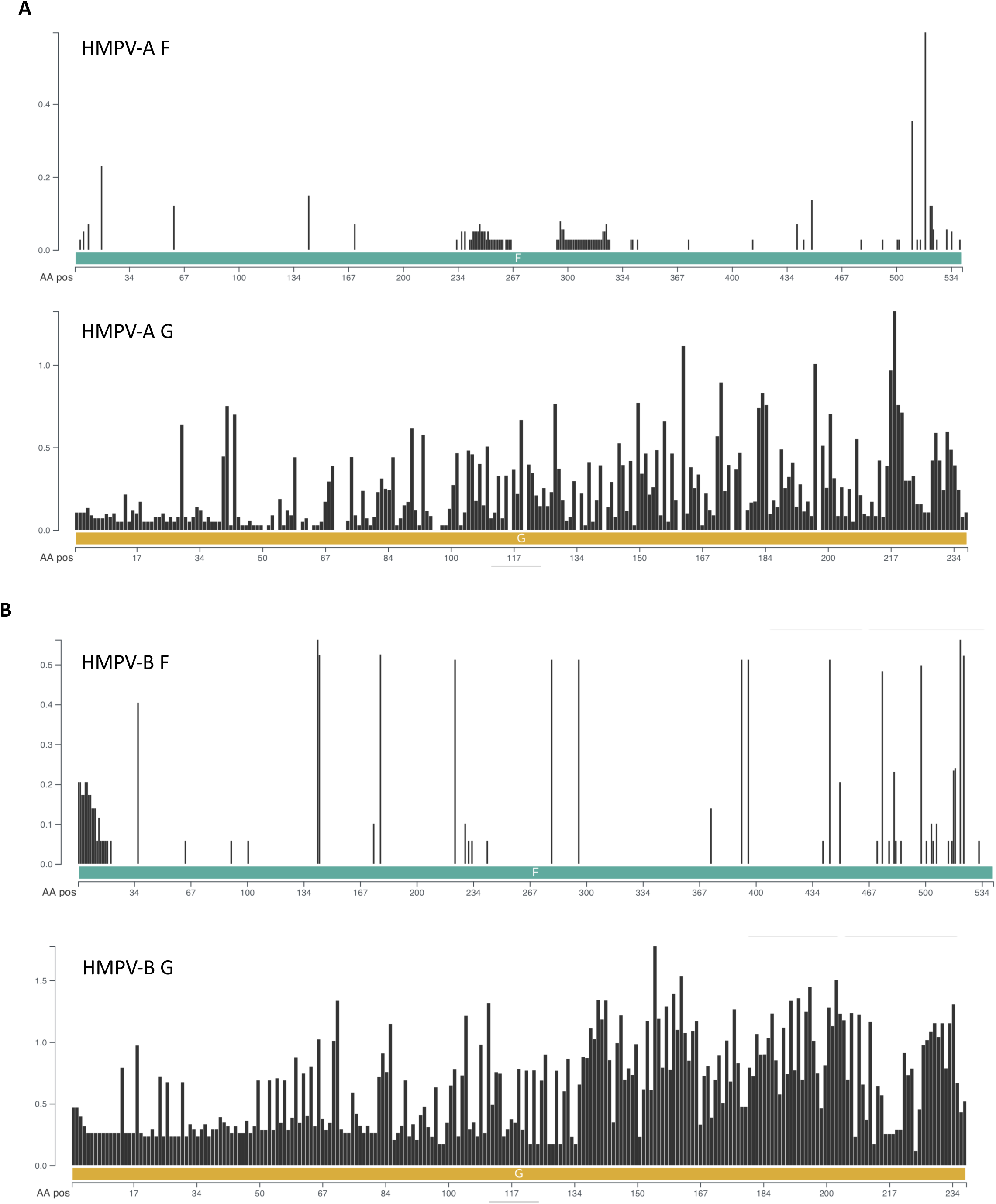
Shannon index for F and G of (A) HMPV-A and (B) HMPV-B.

**Figure S2.**
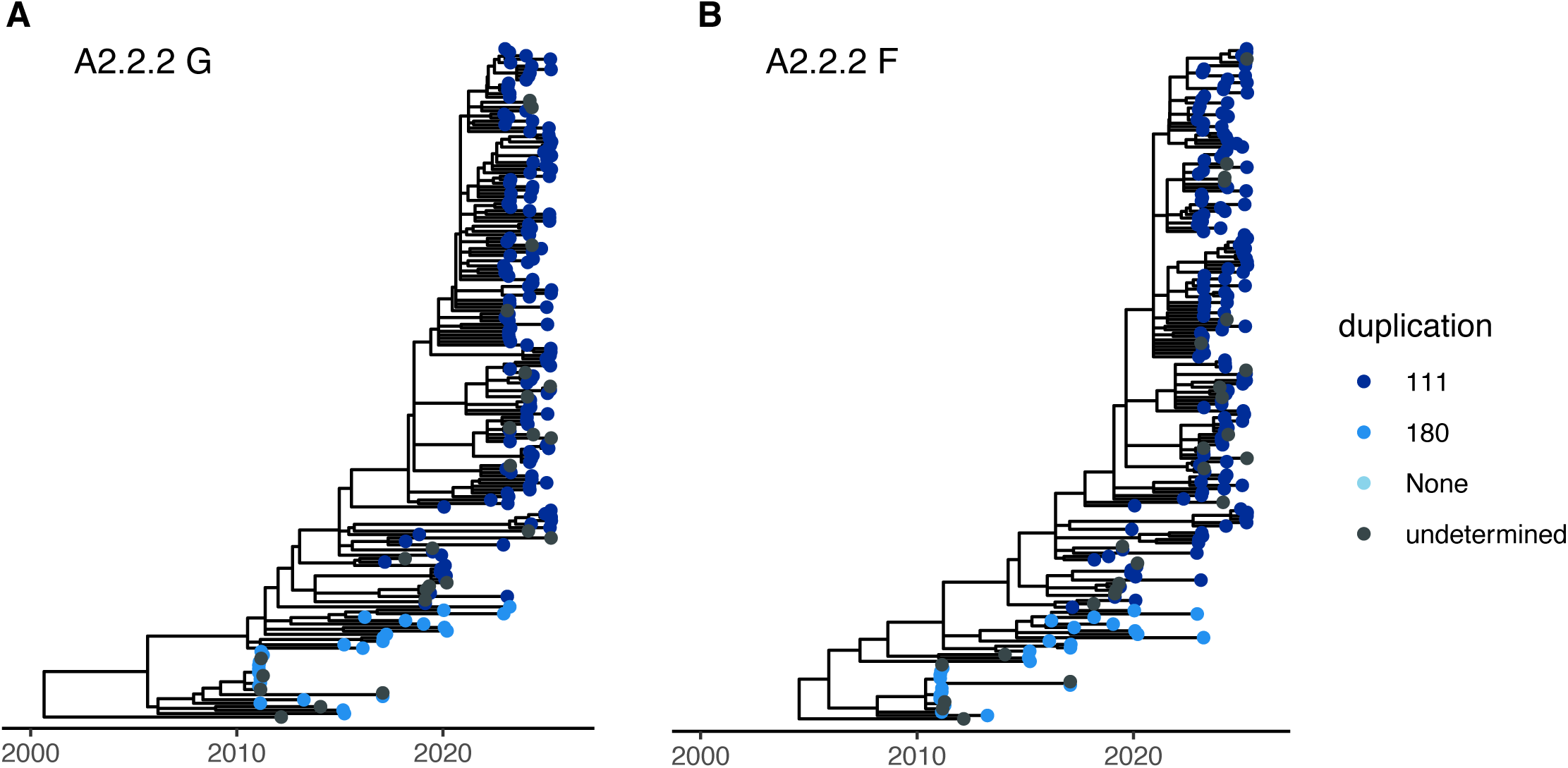
(A) G and (B) F protein phylogeny for A2.2.2 samples. Tips are colored by duplication status.

